# How effective are face coverings in reducing transmission of COVID-19?

**DOI:** 10.1101/2020.12.01.20241992

**Authors:** Joshua F. Robinson, Ioatzin Rios de Anda, Fergus J. Moore, Florence K. A. Gregson, Jonathan P. Reid, Lewis Husain, Richard P. Sear, C. Patrick Royall

## Abstract

In the COVID–19 pandemic, among the more controversial issues is the use of face coverings. To address this we show that the underlying physics ensures particles with diameters ≳1 µm are efficiently filtered out by a simple cotton or surgical mask. For particles in the submicron range the efficiency depends on the material properties of the masks, though generally the filtration efficiency in this regime varies between 30 to 60 % and multi-layered cotton masks are expected to be comparable to surgical masks.

Respiratory droplets are conventionally divided into coarse *droplets* (≳5–10 µm) responsible for *droplet transmission* and *aerosols* (≳ 5–10 µm) responsible for *airborne transmission*. Masks are thus expected to be highly effective at preventing droplet transmission, with their effectiveness limited only by the mask fit, compliance and appropriate usage. By contrast, knowledge of the size distribution of bioaerosols and the likelihood that they contain virus is essential to understanding their effectiveness in preventing airborne transmission. We argue from literature data on SARS-CoV-2 viral loads that the finest aerosols (≳ 1 µm) are unlikely to contain even a single virion in the majority of cases; we thus expect masks to be effective at reducing the risk of airborne transmission in most settings.

## I. INTRODUCTION

The COVID-19 pandemic has brought critically neglected areas of infection control onto the global stage.^1^ Most notably this includes the risk of airborne transmission and the strategies required to mitigate it.^2^ The prevailing view presupposes a binary classification between *large* and *small* droplets (also called aerosols), which respectively transmit disease via the *droplet* and *airborne* routes. This dichotomy originates in research conducted in the 1930s,^3^ and underlies current measures for infection control. Arbitrary threshold diameters (typically 5 to 10 µm) are widely used to distinguish aerosols from large droplets, which have been critiqued by various authors e.g. in Refs. 4–7. In any case, the size distribution of respiratory particles (and their resulting dynamical behaviour) is wide and continuous.

Many outbreaks are suggestive of airborne transmission,^2,8^ aerosolised SARS-CoV-2 virus remains viable under laboratory conditions,^9,10^ viable virus has been cultured from the air surrounding sick patients,^11^ and its predecessor SARS-CoV-1 is considered to have been transmitted by the airborne route.^2,12–15^ Taken together these present a plausible case that the airborne route is significant in transmission of SARS-CoV-2,^2,8,16–19^ although the transmission mechanism by air is complex and many questions remain including the typical viral load of airborne exhaled particles and the required infectious dose. However, absence of evidence is *not* evidence of absence, and so a precautionary approach is justified to improve public safety^1,2,20,21^ similar to that employed to suppress transmission by large droplets or surface contamination where the evidence of transmission is also incomplete.^17,22^

Use of face coverings in public spaces is mandatory or strongly encouraged around the world.^23,24^ Agencies like the Centers For Disease Control and Prevention recommend members of the public wear reusable fabric coverings,^25^ whereas disposable surgical masks are more common in East Asian countries such as China.^26^ Widespread use of surgical masks will inevitably send enormous volumes of plastic waste into the environment, which can take decades (or even centuries) to degrade;^27^ a central question is thus how do fabric coverings perform relative to surgical masks? The efficacy of both types of face coverings is controversial,^21,28,29^ largely because the gold standard of medical evidence is not available for their use; that being said, the current evidence is indicative that they reduce airborne spread.^21,29,30^

It would be unethical to conduct a randomised-placebo controlled trial with high-risk activities such as the spread of SARS-CoV-2, and so we must rely on alternative forms of evidence for the effectiveness of face coverings such as plausibility arguments.^21,31,32^ Here we lay out such a plausibility argument, by combining experimental literature and theoretical calculations. Our goal is to suggest more precisely in which scenarios face coverings are expected to be effective in reducing transmission of SARS-CoV-2. We argue that face coverings do not eliminate the risk of secondary transmissions, especially in high-risk settings such as in hospitals or amongst large crowds, but should significantly reduce the risk in most settings. Moreover, we will suggest criteria where washable cloth masks are expected to be comparable to disposable surgical masks; these may be preferred because they generate less plastic waste.

## II. MASKS ARE PERSONAL AIR FILTERS

A mask or face covering is nothing more or less than an air filter worn on the face. How air filters work and how efficient they are at filtering out particles of differing sizes is reasonably well understood.^35,36^ Droplets and coarser aerosols (with diameter≳1 µm), which are more capable of containing significant viral doses (cf. next section), are more easily filtered because they are less mobile; fine aerosols (0.1 to 1 µm) by contrast are transported around the fibre by the gas flow. We illustrate this schematically in Fig. 1(a).

**FIG. 1.**
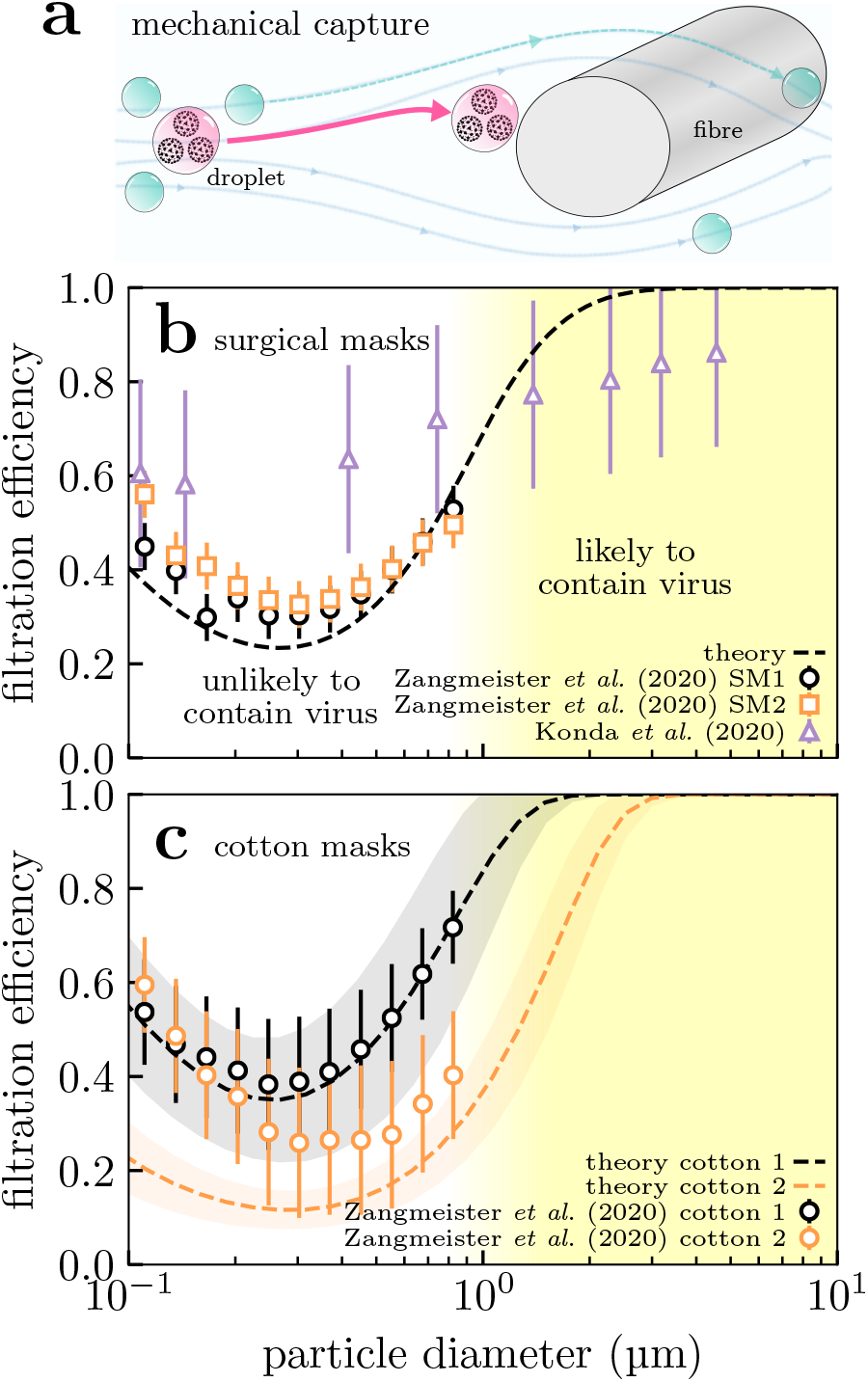
Variation in mask filtration efficiency with incoming particle size. (a) Diagram of capture of viral particles by a fibre within a mask. Larger particles are more easily captured because they are less mobile; smaller aerosols by contrast are transported around the fibre by the gas flow. Larger particles can also carry more virions, and submicron aerosols are unlikely to contain even a single virion (cf. text). The filtration efficiency of perfectly fitting (b) surgical masks and (c) cloth masks formed from 4 identical plain-woven cotton layers are shown as a function of particle size. We show experimental measurements from Refs. 33 and 34 (points) and predictions from our model (lines) which is described elsewhere^35^ and the Supplementary Material. The shaded envelopes around the lines in (c) show the uncertainty in the model predictions, obtained by propagating uncertainties in the geometric parameters given in Ref. 34. We set the velocity of the gas through the mask to 6.3 cm s^−1^ in our calculations for comparison with data from Ref. 34.

In Fig. 1(b) we have plotted both measurements of surgical mask filtering efficiency (symbols) and theoretical calculations (curves). The theoretical calculations involved following the trajectories of particles inserted into the gas flow around fibres, described elsewhere^35^ and in the Supplementary Material (SM). The efficiency is plotted as a function of the particle diameter, because the particle size ultimately determines how hard or easy it is to filter out. The measurements and model agree on the same basic facts:

- Filtering efficiency is essentially 100 % for particles ≳5 µm in diameter or larger.
- However, filtering efficiency is low (30 to 60 %) in the range 0.1 to 1 µm.

These predictions make quantitative the picture we laid out in the preceding paragraph and Fig. 1(a). Both surgical and cotton masks are thus only partially effective at filtering out sub-micrometre aerosols. However, their efficiency rapidly increases as the size increases beyond a micrometre, so masks are generally highly effective in this regime. For reference, SARS-CoV-2 is approximately 0.1 µm in diameter,^37^ so any particle larger than this *can* potentially carry a virus. However, in the next section we will argue that only particles larger than ≳1 µm are *likely* to contain any virus in the majority of cases; thus filtration efficiency in this regime is sufficient to significantly reduce transmission.

Surgical masks are disordered homogeneous arrangements of fibres (cf. Fig. 2(b)), whereas cloth face coverings are typically more ordered (e.g. woven fabric in Fig. 2(a)). More work needs to be done to understand the effect of heterogeneities in cloth masks, but we can obtain some understanding by approximating them as a homogeneous medium^35^ and making a small correction for the effect of inter-yarn pores on collection of submi-crometer particles (SM). In Fig. 1(c) we perform such a calculation for two masks formed from 4 layers of plain-woven cotton fabrics sampled in Ref. 34. Despite considerable variation between different fabrics, the behaviour is more-or-less identical to surgical masks; this is broadly in agreement with the findings of Refs. 33, 34, 38, and 39 and demonstrates that reusable cloth masks can be suitable replacements for disposable surgical masks.

**FIG. 2.**
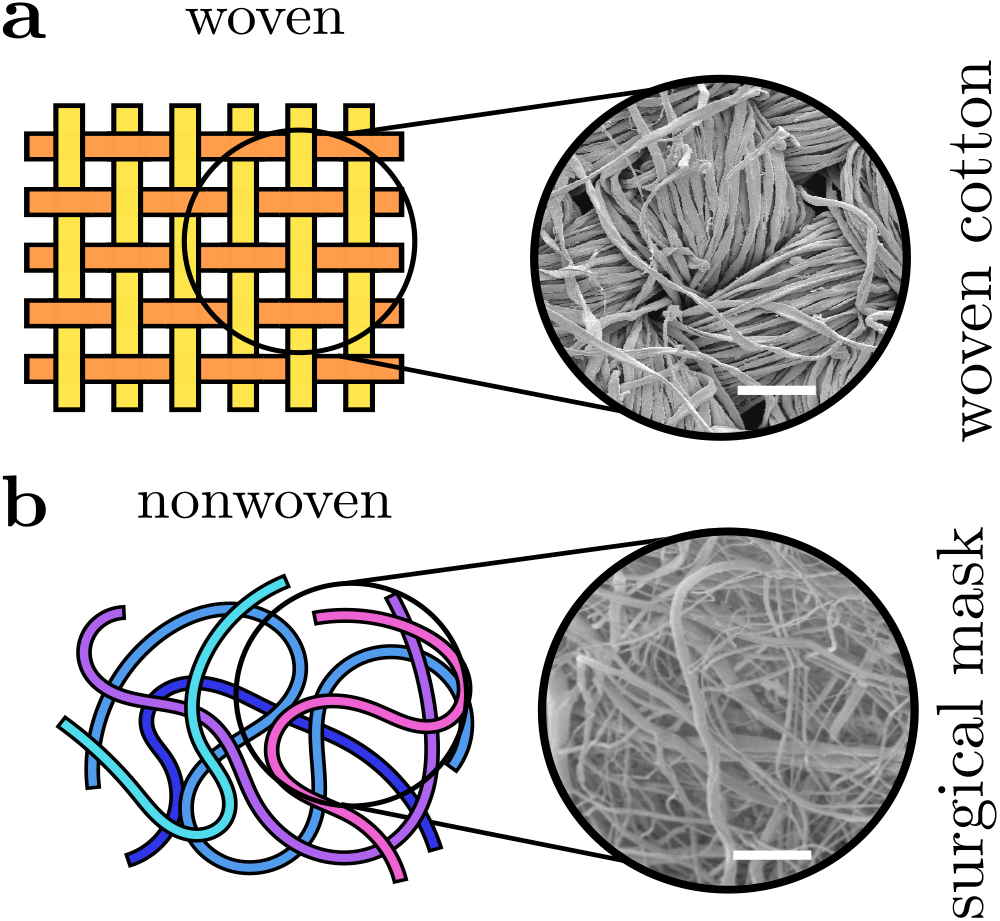
Fabrics are broadly categorised as *knitted* (not shown), *woven* or *non-woven*. (a) Woven fabrics formed by intersecting perpendicular yarns (the “warp” and “weft”). (b) Nonwoven fabrics are formed by entangling fibres through other means, resulting in less ordered arrangements. Scanning electron microscope images of example fabrics show scalebars of (a) 100 µm and (b) 50 µm.

Returning to the mechanism, we briefly discuss the physics underlying this behaviour (more details can be found in Ref. 35). Masks are fundamentally arrays of fibres, see Fig. 2, so air must flow around and between these fibres. Particles a fraction of a micrometre in size have very little inertia and so tend to follow the air flow through the mask faithfully and are not filtered. However, the particle inertia varies with its mass i.e. the cube of its diameter (its volume). This means it rapidly increases with diameter. Beyond around 1 µm in diameter the particles have too much inertia to follow the air as it twists in between the fibres, and so they impact onto the fibres. On microscopic lengthscales most surfaces are attractive, so colliding particles will stick and remain on the fibres.^35^ Because of this basic physics, the filtering efficiency of particles larger than ∼ 5 µm is likely to be limited only by the leakage of air around the mask. For the intermediate range 1 to 5 µm the exact behaviour will depend on the details of the material, but the rapid rise in filtration efficiency with particle size is a robust feature.^35^ Finally, we note that the filtration efficiency increases for capture of the smallest aerosols (≳ 0.3 µm) in Fig. 1(b-c) where capture is enhanced by Brownian motion. We have not focused on this mechanism because such small aerosols are highly unlikely to carry significant doses of virus, and our theory is thus less accurate in this region.

It is useful to compare the effect of wearing a mask with social distancing 1 to 2 m apart. Droplets ~ 20 µm (and smaller) can easily travel several metres,^40–43^ so a perfectly fitting mask would be superior to social distancing for droplets of this size. However, in practice masks are inevitably far from airtight and so many droplets will bypass the mask around the leakage sites (estimated as around 20 % in Ref. 44), and so there is still a case for social distancing even when all parties are masked.

Neither social distancing over 1 to 2 m nor masks are expected to completely mitigate any risk of transmission from sub-micrometre aerosols, because of their long persistence times^6^ and the poor filtration efficiency in this range. The capturing efficiency for particles of diameter 0.1 to 1 µm is only 30 to 60 %, which is a possible cause for concern if these fine aerosols can transmit the virus. Note that respirators (e.g. N95/KN95/FFP2) make use of so-called *electret* fibres which can sustain considerable electrostatic charge;^45,46^ this increases the efficiency in the 0.1 to 1 µm regime. In the next section we will argue that such high-performance filters are unlikely to be needed for most members of the public, and the expected efficacy of face coverings is sufficient for preventing community transmission.

## III. WHICH RESPIRATORY DROPLETS AND AEROSOLS ARE LIKELY TO CONTAIN SIGNIFICANT DOSES OF VIRUS?

To assess the effect of mask wearing we need to know which respiratory droplets and aerosols are likely to carry significant doses of virus. We thus need to know the size distribution of exhaled droplets, and how concentrated the (viable) virus is in droplets produced by shedding in an infected individual.

Instead of an arbitrary size cutoff, expiratory particles can be more meaningfully categorised by their site of origin in the respiratory system. Larger droplets are more likely to be deposited in the respiratory tract,^51–54^ so a large droplet originating deep in the respiratory tract would immediately deposit and be unlikely to escape; as a rule, smaller droplets emerge from lower in the respiratory tract. The majority of droplets produced in the oral cavity vary in size from ∼10 to 1000 µm,^50^ whereas droplets produced in the Larynx and the lower respiratory tract are seen in the range ∼0.1 to 10 µm.^5,49,50,55^ The former presumably contain the majority of virus because volume scales as the diameter cubed^56^, but the latter are expected to be responsible for transmission by the airborne route so they will be our focus.

Fluid particles immediately begin to evaporate upon exhalation^40,41^ and the aerosols will reach their dessicated steady states (which we will refer to as “droplet nuclei”) in less than 1 s.^57^ We use parameterisations of the measured size distributions (i.e. uncorrected for evaporation) reported in Refs. 49 and 50 in our calculations. Their hydrated state would be around a factor of ∼3 larger on exhalation, allowing for coverings to filter aerosols more effectively at the source.^35,41^

Testing for SARS-CoV-2 is primarily performed by detecting the presence of viral RNA in respiratory fluid using reverse-transcriptase polymerase chain reaction (RT-PCR). The distribution of concentrations of viral RNA, the *viral load*, using this technique has been reported in Refs. 47 and 48 which we show in Fig. 3(a). The distribution is extremely broad, spanning around 10–12 orders of magnitude: patients in the upper tail of the distribution (the “superreplicators”) are responsible for hosting most of the virus in circulation, which may be a factor in superspreading events. Each study sampled the viral loads from thousands of patients in all stages of disease progression. These studies primarily involved testing naso-and oropharyngeal samples, which we assume to be representative of *all* respiratory fluids where data is lacking. Finally, we note that there is ample time for evaporation between sample collection and testing in these studies so we assume these concentrations to refer to the dessicated states (the nuclei).

**FIG. 3.**
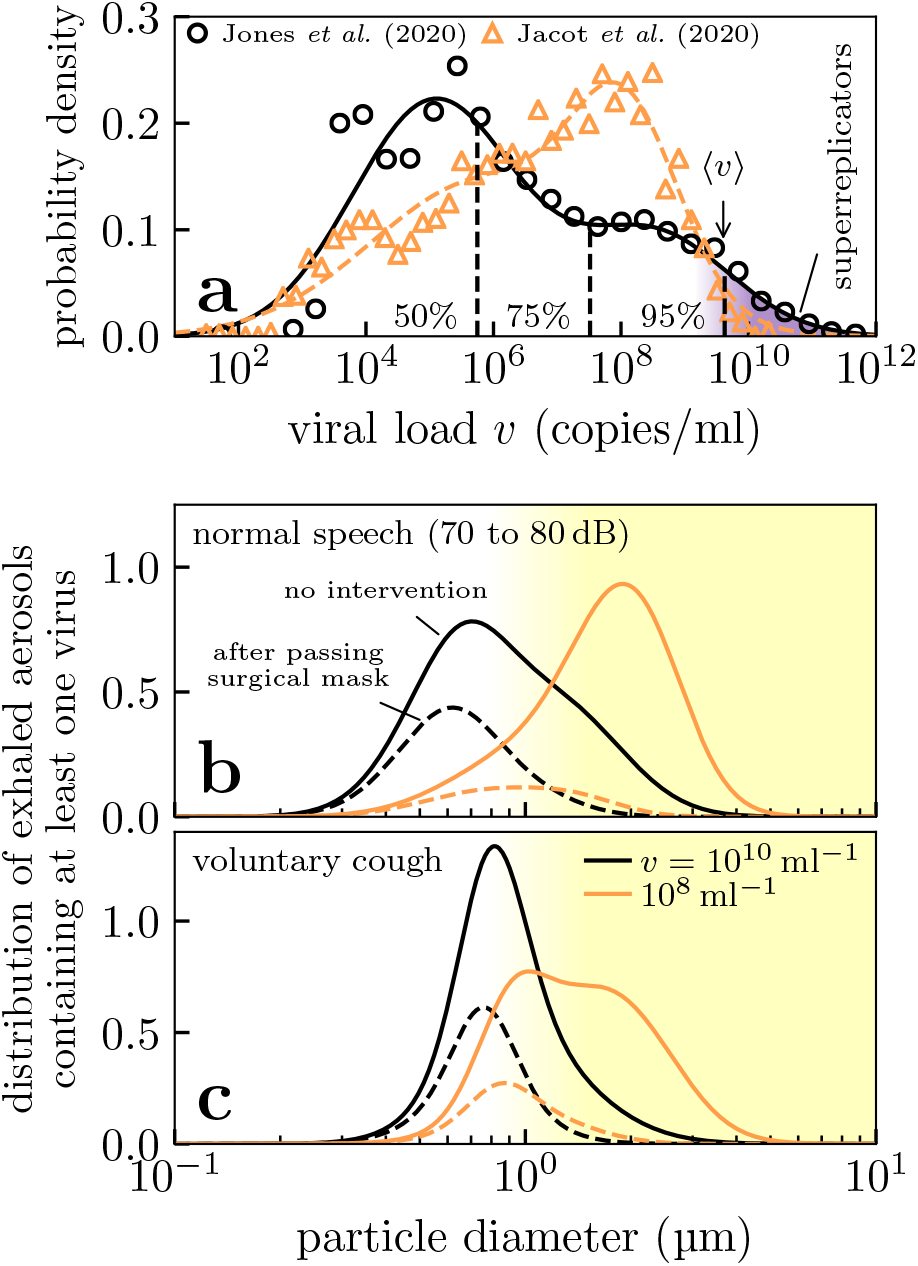
Histograms of key properties of expiratory particles, i.e. normalised so that the area under the curves gives the relative frequency. (a) Distributions of SARS-CoV-2 viral loads in testing from RT-PCR in two studies^47,48^ (points) and our bimodal fits for calculating percentiles (lines). Note the large distribution, and the presence of a tail of patients with extraordinarily large viral loads (shaded purple) corresponding to so-called “superreplicators”. (b-c) Aerosol distributions for virus-laden particles exhaled during speech and voluntary coughing under viral loads typical of the top 25th percentile in (a). We show the distributions of aerosols that contain at least one virus (solid lines) and those that bypass a surgical mass worn by an inhaler (dashed lines); the latter are unnormalised to show the effect of filtering. We calculate the former distributions using data from Refs. 49 and 50 to characterise the exhaled aerosols in healthy patients.

**TABLE 1.** Expected efficacy of surgical masks at filtering virus-laden expiratory particles involved in different transmission modes. These numbers are expected to be similar for multi-layered cloth masks (cf. Fig. 1). The lower bounds were obtained assuming large viral loads of 10^10^ ml^−1^ and 20% mask leakage on inhalation (consistent with Ref. 44) and an estimated 40% leakage for exhalation. The upper bounds were obtained assuming the median viral load of 10^6^ ml^−1^ and perfect fit (no mask leakage).

| Expiratory particles | Fraction filtered |  |  |
| --- | --- | --- | --- |
|  | Inhaler | Exhaler | Both |
| Viral speech aerosols | 47–85 % | 57–99 % | 77–100 % |
| Viral cough aerosols | 49–79 % | 58–99 % | 79–100 % |
| Droplets $\geq 5 \mu\text{m}$ | 80–100 % | 60–100 % | 92–100 % |

Viral load typically peaks around the onset of symptoms^58^ which is also the most contagious stage of disease progression;^8^ we thus expect viral loads from the upper half of the distributions in Fig. 3(a) to be most relevant to disease transmission. The actual number of *viable* virus, as measured from viral plaque assays, in aerosol vs the RT-PCR result is typically only one part in 10^2^ to 10^4^.^10,59^ We thus take the upper limit of viral loads as 10^8^ to 10^10^ ml^*−*1^ instead of 10^10^ to 10^12^ ml^*−*1^.

The probability that an aerosol contains at least one virus depends on its volume and the viral load. By combining this information we can estimate the fraction of particles at any specific size that contains the virus, and thus the variation in the relative number of particles that contain the virus across the size distribution (details of calculation in SM). In Fig. 3(b-c) we show the resulting distributions produced in speech and coughing. For the moderately large viral load of 10^8^ ml^*−*1^ the majority of viral aerosols extend into the micron regime ≳1 µm. Only for extremely large viral loads of 10^10^ ml^*−*1^ do the submicron droplets begin to contain significant numbers of virus. Thus submicron droplets should be unimportant vectors in the majority of people infected with SARS-CoV-2.

## IV. ARE FACE COVERINGS LIKELY TO PREVENT SPREAD IN COMMUNITY SETTINGS?

To understand how masks reduce transmission we need to compare the filtration efficiency of masks in Fig. 1(b) with the droplets that need to be filtered. We do this by showing the distribution of viral droplets that pass through a surgical mask in this with dashed lines in Fig. 3(b-c). We see that masks remove the majority of viral aerosols, leaving primarily those in the submicron regime as expected. Moreover the masks are more effective with decreasing viral loads, so with the exception of “superreplicators” we anticipate masks to be highly effective at reducing transmission rates from the vast majority of people infected with SARS-CoV-2. Where transmission by submicron aerosols is possible, masks can only partially reduce the risk.

While our focus has been on the airborne route, we note here that given face coverings are essentially perfect filters of particles larger than 5 µm in diameter (assuming perfect compliance and appropriate usage) they should provide excellent protection for transmission by the large droplet route and prevent surface contamination. As such, we expect them to provide a net-benefit regardless of whether transmission is dominated by the aerosol or droplet routes.

A key limiting factor for mask efficiency is how much leakage there is around the sides of the mask. For inhalation we expect this to be around 20 %,^44^ though we are not aware of any data on exhalation so we make a conservative (i.e. pessimistic) estimate of 40 % leakage. Note that personal masks can be customised to the individual to improve the fit, so these are upper bounds. Accounting for mask leakage we estimate the total fraction of droplets and aerosols filtered in table I when masks are worn by either the (susceptible) inhaler, the (infected) exhaler or both people. Even with greater leakage masking the exhaler is generally more effective at preventing airborne transmission than masking the inhaler, because aerosols are larger (and thus filtration is more effective) before the aerosols evaporate into their dessicated states. We have underestimated the protection from large droplets whilst masking the exhaler because the mask would deflect and remove momentum from the expiratory jets;^60,61^ taking this into account, masking the exhaler (“source control”) is expected to offer more protection than masking the inhaler. In all cases masking both people is more effective as this partially makes up for deficiencies in leakage and the worse filtration in the submicron regime.

## V. DISCUSSION

The SARS-CoV-2 pandemic has exposed critical knowledge gaps in infection control and prevention.^1,62^ We believe these gaps necessitate cross-disciplinary collaborations on matters of public policy, and in this spirit we have presented a physical scientists’ perspective on the effectiveness of face coverings.

Both masks made from simple cotton fabrics, and surgical masks, are predicted to reduce transmission of respiratory viruses. Cloth masks can be washed and reused making them preferable to disposable surgical masks because they generate less plastic waste which is better for the environment. As masks are cheap, and wearing a mask is a relatively minor inconvenience compared to contracting SARS-CoV-2, recommending mask use is a simple way to reduce transmission. The extent of the reduction depends to some degree on whether transmission is via large droplets or aerosols, however we have argued that in all cases masks are likely to significantly reduce the risk from viral particulates. Due to the inevitable poor fit of these masks, filtration is never 100 % effective for any transmission mode, but they filter larger droplets much more efficiently than smaller ones.

Transmission of respiratory viruses is complex and poorly understood, so more data is needed. We need either direct data on transmission rates as a function of conditions, with and without masks, and a much better idea of the infectivity of aerosolised virus including the required dose for infection Both of these will be challenging but both are possible. However, the basic physics of filtration tells us about how capture varies with droplet size.

Both masks and social distancing reduce the exposed dose by some percentage, dependent on how big the droplets are that are involved in transmission. It now seems well established that with SARS-CoV-2 some infected people have viral loads thousands or millions of times higher than others.^47,48^ Thus a say 50 % reduction in dose due to mask wearing corresponds to very different absolute reductions in dose from infected people with high and low viral loads. As typically the viral load of an infectious person will not be known, other forms of interventions may be warranted in addition to masking.

## Data Availability

All underlying code is available at:
https://github.com/tranqui/maskflow

## ACKNOWLEDGMENTS

The authors wish to thank Kate Oliver for helpful discussions on textiles, Patrick Warren for guidance on LB simulations, Mahesh Bandi for making us aware of his ingenious use of a candyfloss maker, and Mike Allen, Jens Eggers, and Daan Frenkel for helpful discussions We gratefully acknowledge Daniel Bonn, Patrick Charbonneau, K. K. Cheng, Rosie Dalzell, Tanniemola Liverpool, John Russo and Hajime Tanaka for providing valuable comments on this work.

JFR, JPR and CPR wish to thank the Bristol Aerosol COVID-19 group for valuable discussions and feedback on this work. JFR would like to thank Kirsty Wynne for assistance in debugging the code used in the theoretical calculations. The authors would like to thank Judith Mantell and Jean-Charles Eloi of the Wolfson Bioimaging Facility and the Chemical Imaging Facility (EPSRC Grant “Atoms to Applications”, EP/K035746/1), respectively, for the SEM images and assistance in this work.

