## Supplementary Material for "How effective are face coverings in reducing transmission of COVID-19?"

### I. THEORETICAL MODEL FOR FILTRATION

Here, we briefly summarise the theoretical model used to obtain the theoretical filtration curves in the main text. We have provided a full account of this model in Ref. 1, which goes through this theory in more detail for an audience of fluid mechanics specialists. The theory is a development in filtration theory, which is widely used in the treatment of air filters. Our developments have extended its applicability to fabrics, which enabled the agreement between theory and experiments shown in Fig. 1 in the main text.

#### A. Particle motion

We consider particles of radius $a_p$ moving at velocity $\mathbf{v}$ while being transported by a gas flow field $\mathbf{u}$. To model the gas flow, we treat the mask as a system of infinitely long cylinders of radius $a_f$ occupying a volume fraction $\alpha$ inside the mask. Assuming the effective neighbourhood around each fibre can be modelled by the co-axial region with distance from its centre $r \in [a_f, a_f/\sqrt{\alpha}]$ and Stokes flow yields the Kuwabara flow field for $\mathbf{u}$ [1, 2]. We write the characteristic flow speed through the mask as $U_0$, which typically takes on values of $U_0 \sim \mathcal{O}(1\,\mathrm{cm\,s^{-1}})$ for flows through a face covering [1].

Next we insert the particles into the gas flow and follow their trajectories. Assuming Stokes flow, the particle equation of motion reads

$$m_p \frac{d\mathbf{v}}{dt} = -\frac{\mathbf{v} - \mathbf{u}}{B} \tag{1}$$

where $m_p$ is the particle mass. The term on the right hand side is the Stokes' drag. In this term $B = C/6\pi\mu a_p$ is the particle mobility, with $\mu$ the dynamic viscosity of air and $C$ the Cunningham slip correction factor. We have assumed that the particle interacts with the flow field as a point particle so that: (a) the flow field $\mathbf{u}$ is unperturbed by the presence of the particle and (b) the Stokes drag couples only to the particle's centre of mass.

#### B. Filtration efficiency

To understand the filtering capacity of a single fibre, we consider the flow around an infinitely long cylinder aligned perpendicular to the direction of flow. We define the single-fibre efficiency $\eta$ as the fraction of particle trajectories that terminate on the fibre surface. We further assume that far from the fibre the particles are:

1. Aligned with the gas flow field, which is uniform before being disturbed by the fibre.
2. The particles are randomly positioned in the gas flow.

The single-fibre efficiency can then be expressed as $\eta = \lambda/W$ where $W$ is the width of the mask (approximately the width of the face), which introduces $\lambda$ as the width of the *collection window*. If the particle is initially placed inside this window then it will ultimately terminate on the surface of the fibre, otherwise it will bypass the fibre.

In the main text we argued that the main collection mechanism for masks arises from particle inertia (also called "impaction"), and we indicated that Brownian motion (diffusion) acts as a secondary mechanism for the smallest aerosols. We respectively write the single-fibre efficiency for these two mechanisms as $\lambda_I$ and $\lambda_D$. These mechanisms act in distinct size regimes, so their combined efficiency is well-approximated as.

$$\lambda \simeq \lambda_I + \lambda_D. \tag{2}$$

We numerically determined the inertial term $\lambda_I$ by repeatedly inserting particles into the gas flow field at varying initial conditions and integrated (1). As diffusion is the less important mechanism in this work, we have simply made use of the standard relation [3]

$$\frac{\lambda_D}{2a_f} = \frac{2.9}{(K\mathrm{Pe}^2)^{1/3}} + \frac{0.624}{\mathrm{Pe}} + \frac{1.24R^{2/3}}{\sqrt{K\mathrm{Pe}}}, \tag{3}$$

where $R = a_p/a_f$, Pe is the Péclet number and $K$ is the hydrodynamic factor emerging from the Kuwabara flow field [2].

#### C. Filtration efficiency of masks

We consider the mask as an assembly of perfectly cylindrical fibres aligned perpendicular to the flow and randomly distributed with uniform probability. We find that the *penetration*, the fraction of particles that pass through the mask without being filtered, is [1]

$$P = \exp\left(-\frac{L}{\xi}\right), \tag{4a}$$

with penetration length

$$\xi = \frac{(1-\alpha)\pi}{4\alpha\overline{\lambda}} \int_0^\infty d_f^2\, p(d_f)\, dd_f, \tag{4b}$$

and effective collection window

$$\overline{\lambda} = \int_0^\infty \lambda(d_f)\, p(d_f)\, dd_f. \tag{4c}$$

where $L$ is the thickness of the mask, $\alpha$ is the volume fraction occupied by the fibres and $p(d_f)$ gives the distribution of diameters in the sample which are well-described by a log-normal distribution i.e. $\ln(d_f/\mu\mathrm{m}) \sim \mathcal{N}(\mu, \sigma^2)$. We have previously determined the parameters characterising fibres in surgical masks and cotton fabrics from scanning electron microscope images [1].

Cotton fibres have $\mu = 2.68$ (corresponding to a modal fibre diameter of 14.6 µm) and $\sigma = 0.34$ [1]. Woven fabrics are heterogeneous: the fibres are twisted into yarns which are then woven into their final structure as shown in Fig. 2(a). To treat this heterogeneity we start by calculating $\lambda$ for an effective homogeneous system, and then we correct for the fact that most of the flow occurs through the inter-yarn pores where the fibres are less dense. We calculate the average thickness and volume fraction for the plain-woven cotton fabrics considered in Ref. 4 from their stated values of areal density, yarn thicknesses and yarn densities. To take this average we assume the yarns are cylindrical so that the maximum thickness is thus the sum of the warp and weft yarn diameters; the average thickness is thus less than this maximum thickness because of the pores and the cylindrical shape of the yarns. The average volume fraction can then be estimated by combining this quantity with the areal mass and the bulk density of cotton. For the two fabrics in Fig. 1(c) we find average thicknesses of $L = 0.16$ mm and 0.24 mm and average volume fractions of $\alpha = 0.48$ and 0.26.

The flow speed is essentially zero inside the yarn, so most of the flow goes through the inter-yarn pores. Consequently, compared to flow through a homogeneous material: (i) the effective fibre density will be reduced and (ii) the typical flow speed will be increased. Effect (i) generically lowers the collection efficiency as there are fewer fibres to collect particles, whereas the effect of (ii) depends on the collection mechanism. Collection by inertia (impaction) is enhanced by increasing the flow speed, opposing the effect from an effectively reduced fibre density. After cancellation we thus expect the resulting change in efficiency to be small, and so we do not correct this collection mechanism. However, the efficiency of collection by diffusion decreases with increasing flow speed, reinforcing effect (i), which is potentially significant. We correct $\lambda_D$ by inserting a locally increased flow speed into the Péclet number entering (3). We estimate the fabric permeability $k$ (the area fraction of pores) from the yarn parameters given in Ref. 4, and from this we obtain the flow speed inside the pores as $U_0 \to U_0/k$. No fitting was required to match the experiments of Ref. 4 because all parameters are known.

For surgical masks we found the inner side of the mask was composed of fine fibres with $0.75 \lesssim \mu \lesssim 1$ and $0.35 \lesssim \sigma \lesssim 0.45$, and on the outer side they were coarser with $3 \lesssim \mu \lesssim 3.2$ and $0.07 \lesssim \sigma \lesssim 0.1$ [1]. We model this as two layers of thickness $L_1$ and $L_2$ with total thickness $L_1 + L_2 = L \simeq 0.8$ mm determined from optical microscopy [1]. We set $\mu_1 = 0.9$ (mode 2.5 µm), $\mu_2 = 3$ (mode 20 µm), $\sigma_1 = 0.4$, $\sigma_2 = 0.1$ and the volume fraction as $\alpha = 0.075$ in both layers (determined previously in Ref. 1 by weighing samples). We fit the total penetration to the experimental data to find the relative thicknesses of each layer $L_1$ and $L_2$, with the fit performed while their combined thickness was kept fixed. Surgical masks are nonwoven (and thus homogeneous) so we did not need to correct for changes in collection efficiency due to large inter-yarn pores.

### II. EXHALED AEROSOL DISTRIBUTIONS CONTAINING AT LEAST ONE VIRUS

The average number of virions in a spherical homogeneous particle of diameter $d_p = 2a_p$ with viral concentration $v$ is

$$\langle n_v \rangle = v \frac{\pi d_p^3}{6}.$$

Assuming the Poisson distribution, the probability that a particle of this size contains at least one virion is

$$p(v|d_p) = 1 - e^{-\langle n_v \rangle}.$$

This probability is negligible for $d_p \lesssim 1\,\mu\mathrm{m}$ for $v \ll 10^{10}\,\mathrm{ml}^{-1}$, as we stressed throughout the main text. Application of Bayes' theorem gives the probability distribution for viral aerosols as

$$p(d_p|v) \propto p(v|d_p)p(d_p).$$

with the proportionality constant ensuring that the final distribution is normalised. The quantity $p(d_p)$ is the distribution of *all* exhaled aerosols (i.e. viral or otherwise), which we take directly from the fits to experimental data in Refs. 5 and 6. Multiplying these distributions by $d_p$ gives the probabilities normalised with $\ln d_p$ instead of $d_p$; we did this in Fig. 3 so that the area under the curve corresponds to the frequency.

---


[1] Robinson JF, Rios de Anda I, Moore FJ, Reid JP, Sear RP, Royall CP. Efficacy of face coverings in reducing transmission of COVID-19: calculations based on models of droplet capture; 2020. Preprint: arXiv 2008.04995 awaiting journal submission. Available from: `https://arxiv.org/abs/2008.04995`.

[2] Kuwabara S. The Forces Experienced by Randomly Distributed Parallel Circular Cylinders or Spheres in a Viscous Flow at Small Reynolds Numbers. J Phys Soc Jpn. 1959 Apr;14(4):527–532.

[3] Stechkina IB, Fuchs NA. Studies on Fibrous Aerosol Filters—I. Calculation of Diffusional Deposition of Aerosols in Fibrous Filters. Ann Occup Hyg. 1966 Apr;.

[4] Zangmeister CD, Radney JG, Vicenzi EP, Weaver JL. Filtration Efficiencies of Nanoscale Aerosol by Cloth Mask Materials Used to Slow the Spread of SARS-CoV-2. ACS Nano. 2020 Jul;14(7):9188–9200.

[5] Gregson FKA, Watson NA, Orton CM, Haddrell AE, McCarthy LP, Finnie TJR, et al. Comparing the Respirable Aerosol Concentrations and Particle Size Distributions Generated by Singing, Speaking and Breathing; 2020.

[6] Johnson GR, Morawska L, Ristovski ZD, Hargreaves M, Mengersen K, Chao CYH, et al. Modality of Human Expired Aerosol Size Distributions. J Aerosol Sci. 2011 Dec;42(12):839–851.
